# Development and validation of a Trans-Ancestry polygenic risk score for Type 1 Diabetes

**DOI:** 10.1101/2025.06.09.25329166

**Authors:** Basile Jumentier, HuiQi Qu, Tianyuan Lu, Kai Liu, Erica L. Kleinbrink, Kathleen Klein, Wiame Belbellaj, Isabel Gamache, Lauric Ferrat, Guillaume Butler-Laporte, Yangxi Li, Hakon Hakonarson, Wei Wu, Constantin Polychronakos, Celia M.T. Greenwood, Despoina Manousaki

## Abstract

**Objectives:** The high heritability of type 1 diabetes has enabled the development of polygenic risk scores (PRS) as disease risk screening tools. PRS can identify individuals at the highest genetic risk in a population, who can benefit from autoantibody and metabolic surveillance, to avoid ketoacidosis at diagnosis and access preventive therapies. However, PRS for type 1 diabetes developed from European data perform less well in non-European ancestries. We aimed to develop a PRS with comparable performance among different ancestries.

**Methods:** Using a the PRS-CSx method, and data from large European, East-Asian, African-American and Hispanic type 1 diabetes GWAS (N_total_cases_=29,469), we developed a trans-ancestry PRS (TA-PS), combining a non-*HLA* component incorporating over a million variants, with the *HLA* component of a published European PRS (GRS2x). We tested the performance of the PRS using AUROC, sensitivity and specificity in a multi-ancestry T1D case-control cohort (N_total_= 4,657; N_non-European_=556) from Montreal, Canada. We validated our results in two independent T1D case-control cohorts (CHOP-CAG and GRACE) and two population-based cohorts (All of Us and UK Biobank).

**Results:** In our multi-ancestry Montreal-based cohort, TA-PS showed an AUROC of 0.89 which was significantly higher from the AUROC of 0.85 of GRS2x. At a 90th percentile cut-off, in African-Americans, the sensitivity of GRS2x was 0.32, compared to 0.56 in Europeans. For TA-PS, we obtained overall better sensitivities, ranging from 0.71 in Europeans to 0.77 in South Asians. TA-PS demonstrated slightly lower albeit acceptable specificity compared to that of GRS2x (> 0.83 across all ancestries). These results were validated in the four independent cohorts.

**Conclusion:** We developed a trans-ancestry PRS that outperformed the European-based GRS2x. Importantly, TA-PS provides a comparable prediction in various ancestries, which supports its use in population-wide screening programs.

**Research in context:** *What is already known about this subject?:*

- Polygenic risk scores (PRS) for type 1 diabetes are primarily developed using data from individuals of European ancestry.
- The widely used, European-based GRS2 score shows reduced performance in non-European populations, particularly among individuals of African descent.
- There are concerns regarding the equity of genetic risk prediction in population-based screening programs for T1D.

*What is the key question?:*

- Can a trans-ancestry PRS provide accurate and equitable type 1 diabetes risk prediction across ancestries?

*What are the new findings?:*

- A new trans-ancestry score, TA-PS, was developed by integrating an optimized non-*HLA* PRS to GRS2.
- Compared to GRS2, in multi ancestry case-control and population-based cohorts, TA-PS improves sensitivity across all ancestry groups while maintaining high specificity.

*How might this impact on clinical practice in the foreseeable future?:*

- TA-PS could provide equitable genetic risk stratification in population-wide screening programs for type 1 diabetes.

## Introduction

Type 1 diabetes is a chronic disease caused by the autoimmune destruction of pancreatic β cells, which are responsible for insulin production. The prevalence of type 1 diabetes is strongly influenced by ancestry. Notably, its prevalence is 4 in 1000 in White Europeans but generally lower in other ancestries [1]. Family studies suggested that 50% of type 1 diabetes risk is explained by genetics [2]. The majority of this genetic risk can be explained by the variation of a few loci within the *HLA* class 2 region (DR-DQ) but also by about one hundred non-*HLA* single nucleotide polymorphisms (SNPs) including SNPs in the *INS* and *PTPN22* genes [3]. The high heritability and the identification of a large number of loci associated with type 1 diabetes has allowed the development of powerful screening tools based on polygenic risk scores (PRS) [4–7]. Among these, a score named GRS2 [8], developed from European data and comprising 67 SNPs, including 32 non-*HLA* SNPs and *HLA* interactions, presents the highest predictive performance with an Area Under the Receiver Operating Curve (AUROC) of 0.92 in a European case/control cohort (N = 15728, cases = 6481). It has been shown that within the highest 10% of the GRS2 distribution in a general European population, up to 84% of cases of incident type 1 diabetes cases can be identified, which makes GRS2 a useful tool for type 1 diabetes population-wide screening programs [9]. However, as expected, GRS2’s performance varies widely depending on the ancestry. For instance, in a cohort of African-Americans (N = 1534, cases = 168), its AUROC was 0.81 [10]. Furthermore, GRS2 distributions differ depending on the ancestry [1,11], which complicates the choice of a threshold in screening programs, especially in regions or countries with multi-ancestral populations[7,9]. Against this backdrop, the aim of this study was first, to develop a PRS for type 1 diabetes with comparable performance across ancestries and, second, to identify a uniform threshold of this PRS to define high risk for application in future screening programs.

To do this, we focused on optimizing the non-*HLA* component of GRS2. Leveraging large and recent genome-wide association studies (GWAS), on European [12], East Asian [13], African-American and Hispanic [14,15] populations, and machine learning methods for trans-ancestry PRS development [16–18], we built a series of trans-ancestry PRS. The model selection was done in a multi-ancestry cohort (N_cases_ = 2416, N_controls_ = 2241), assembling existing case-control collections and newly genotyped samples. Our best performing trans-ancestry PRS (named TA-PS) was then validated in population-based or case-cohorts assembling 3230 cases from multiple ancestries. For our second aim, the same datasets were used to choose a threshold of the TA-PS with the optimal combination of sensitivity and specificity.

## Materials and Methods

### Training of ancestry-specific and trans-ancestral PRSs

To develop our trans-ancestry scores, we optimized a previous European-based score, GRS2x. GRS2x [19] is an extension of the 67 SNP-score GRS2 [8], which is adapted to data imputed with TOPMed (R2 and R3) or whole genome sequencing (WGS) data. GRS2x can be split into 3 components: the *HLA* interaction part (GRS2x_Int_) containing 14 *HLA* DR/DQ SNPs as well as interaction terms, the *HLA* part (GRS2x_HLA_) containing 21 non-DR/DQ *HLA* SNPs and the non-*HLA* part (GRS2x_NonHLA_) containing 32 non-*HLA* SNPs.

For the trans-ancestry optimization of GRS2x, we modified the GRS2x’s non-*HLA* part using the published methods PRS-CSx [17] and JointPRS [16]. For comparison purposes, we also used LDpred2 to optimize the GRS2x’s non-*HLA* part for Europeans [18]. To ensure absence of *HLA* overlap in the non-*HLA* trans-ancestry PRS, we excluded the MHC region (hg19: chr6:28,477,797-33,448,354) and a 1Mb LD window around it [20].

Both PRS-CSx and JointPRS algorithms produce a range of ancestry-specific (AS) or trans-ancestral (TA) scores with a varying shrinkage parameter (phi) based on GWAS summary statistics (**Supplementary Table 1**) from large European and smaller ancestry-specific GWAS and ancestry-matched LD reference panels. We used the largest published European GWAS [12] (N_EUR_cases/controls_ =18942/501638) and East Asian GWAS [13] (N_EAS_cases/controls_ = 1219/132032). For African-American and Hispanic ancestries, we meta-analyzed two recent GWAS (N_AFR_Michalek_cases/controls_ = 409/482, N_AFR_Verma_cases/controls_ = 6451/115861; N_AMR_Michalek_cases/controls_ = 153/158, N_AMR_Verma_cases/controls_ = 2295/55134) [14,15] using METAL [21].

Using the above methods, we generated 35 PRS for T1D (**Supplemental Note 1**): using PRS-CSx, we built 6 ancestry-specific PRS per ancestry (6 phi tested for AFR, AMR, EAS and EUR) and 6 trans-ancestral PRS (6 phi tested); using JointPRS, we generated one ancestry-specific PRS per ancestry (for AFR, AMR, EAS, EUR); finally, we used LDpred2 to develop an optimized European PRS for comparison purposes.

### Description of test cohorts

In order to test the newly developed trans-ancestry PRS against GRS2x, we assembled a multi-ancestry cohort of T1D cases and controls. This cohort includes previously published Montreal-based cohorts such as: CaCo (n_cases_ = 514; n_controls_ = 2027), MTL (n_cases_ = 482) [22], and newly genotyped T1D cases as part of this project (n_cases_ = 1086), and from the Montreal Children’s Hospital T1D Biobank (n_cases_ = 334). Using principal component analysis, we paired the cases of the latter three cohorts to ancestrally-matched controls from the 1000 Genomes Project [23] (1KGP) database (n_controls_ = 214) (**Supplementary Table 2**) using the R package ‘PCAmatchR’ (version 0.3.3). In total, we assembled 2416 T1D cases and 2241 controls from 5 different populations: African (N = 139), Hispanic (N = 236), East Asian (N = 56), European (N = 4101) and South Asian (N = 125), determined by principal component analysis using the 1KGP reference. Type 1 diabetes diagnosis was defined according to clinical criteria in all cohorts, except in the Montreal Children’s T1D Biobank, where confirmation with autoantibodies was available. All genotypes were imputed using TOPMed (R3) [24] except for controls from 1KG, where WGS data were available. As part of the post-imputation quality control, we applied filters for minor allele frequency (MAF > 0.001) and imputation quality (info-score > 0.4).

### Computation of the PRS

To compute all 35 newly developed PRSs and the GRS2x (GRS2x_HLA_ and GRS2x_NonHLA_ terms) from our imputed data, we used plink1.9 and its "--score" command. For GRS2x_Int_, we used the toolkit developed by Sharp et al available on Github (USF-HII/*HLA*-prs-toolkit). This toolkit allows generating scores based on SNPs (here the 14 DR/DQ SNPs) for type 1 diabetes where the epistasis of the *HLA* genes (gene versus gene interaction) is modelled using SNPs in strong linkage disequilibrium with the *HLA* haplotypes.

In order to combine the 35 new non-*HLA* PRS with the *HLA* and the *HLA* interaction part of the GRS2x, we applied different approaches. First, since the 35 non-*HLA* scores did not have the same mean and standard deviation as the GRS2x, we scaled each of the *HLA* and non-*HLA* components to have zero mean and unit variance (*scale* function in R). For instance, a trans ancestral non-*HLA* PRS generated with PRS-CSx represented the sum of scale(TA) + scale(GRS2x_Int_ + GRS2x_HLA_).

Second, to further optimize the combination of the different scores, we estimated coefficients for the 3 terms of our models (non-*HLA*, interaction and *HLA*). These coefficients were estimated only from European data (N_cases_ = 2193; N_controls_ = 1908) using logistic regression: (T1D_case_status_ ∼ TA + GRS2x_Int_ + GRS2x_HLA_). The coefficients were then extracted from this regression model (“a” for non-*HLA*, “b” for GRS2x_Int_ and “c” for GRS2_HLA_) and applied as follows: fit = a * non-*HLA* + b * GRS2x_Int_ + c * GRS2x_HLA_. The coefficients a, b and c were estimated using two transformations of each term with either scale or MinMax (**Supplementary Table 3**). The MinMax transformation scales data to a specified range, here between 0 and 1, by subtracting the minimum value and dividing by the range of the data.

### Testing of the PRSs

To test the prediction performance across various ancestries of all generated PRS, and of the GRS2x, we calculated the area under the ROC curve (AUROC + 95% CI). We used the Delong test to compare the AUROC between two PRS within the same ancestry. This led to the selection of the best performing PRS for each ancestry, and across all ancestries.

To evaluate the optimal threshold of the best performing PRS, we calculated, across all ancestral datasets and in our multi-ancestry cohort, sensitivity and specificity metrics at different thresholds based on European control data. We considered the following thresholds: top 5%, 10%, 15%, 20% and 25% of the distribution (corresponding to the 95^th^, 90^th^,85^th^, 80^th^ and 75^th^ centile). For each threshold, we also calculated the F1-score metric, which corresponds to the harmonic mean of precision and recall. Confidence intervals for these different metrics were estimated by bootstrap (n_iter_ = 1000) and the comparison of these values was carried out with a Z-test.

### Validation in independent cohorts

We then tested the performance of the various PRS in independent case/control or population-based cohorts of various ancestries (**Supplementary Table 4**). Specifically, in the multi-ancestry All of Us Biobank (AoU, N = 89,699; N_cases_ = 1084; N_controls_ = 88,635), in a case-control multi-ancestry cohort from the Children’s Hospital of Philadelphia (CHOP-CAG, N = 6498; N_cases_ = 1094; N_controls_ = 5404), in the UK Biobank (UKB, N = 427,847; N_cases_ = 778; N_controls_ = 427,069) and in a Chinese case/control cohort across multiple pediatric centers named Genetic Risk Assessment for Chinese Eaglet-T1D (GRACE, N = 597; N_cases_ = 294; N_controls_ = 303). AoU policy prohibits conducting stratified analyses on ancestry if the number of cases is less than 20, and we applied this approach to all cohorts. The definition of cases and controls for each cohort is available in **Supplementary Note 2.** Ethics approvals for use of samples of all the above studies was obtained by the respective ethics committees.

For each cohort, we estimated the AUROCs of the different PRS and the other three metrics at the following thresholds: 5%, 10%, 15%, 20% and 25%. The thresholds were estimated only on the controls for the case/control cohorts while for the population cohorts the entire datasets were used.

The flowchart with the analytical steps of our study appears in **Figure 1**.

**Figure 1.**
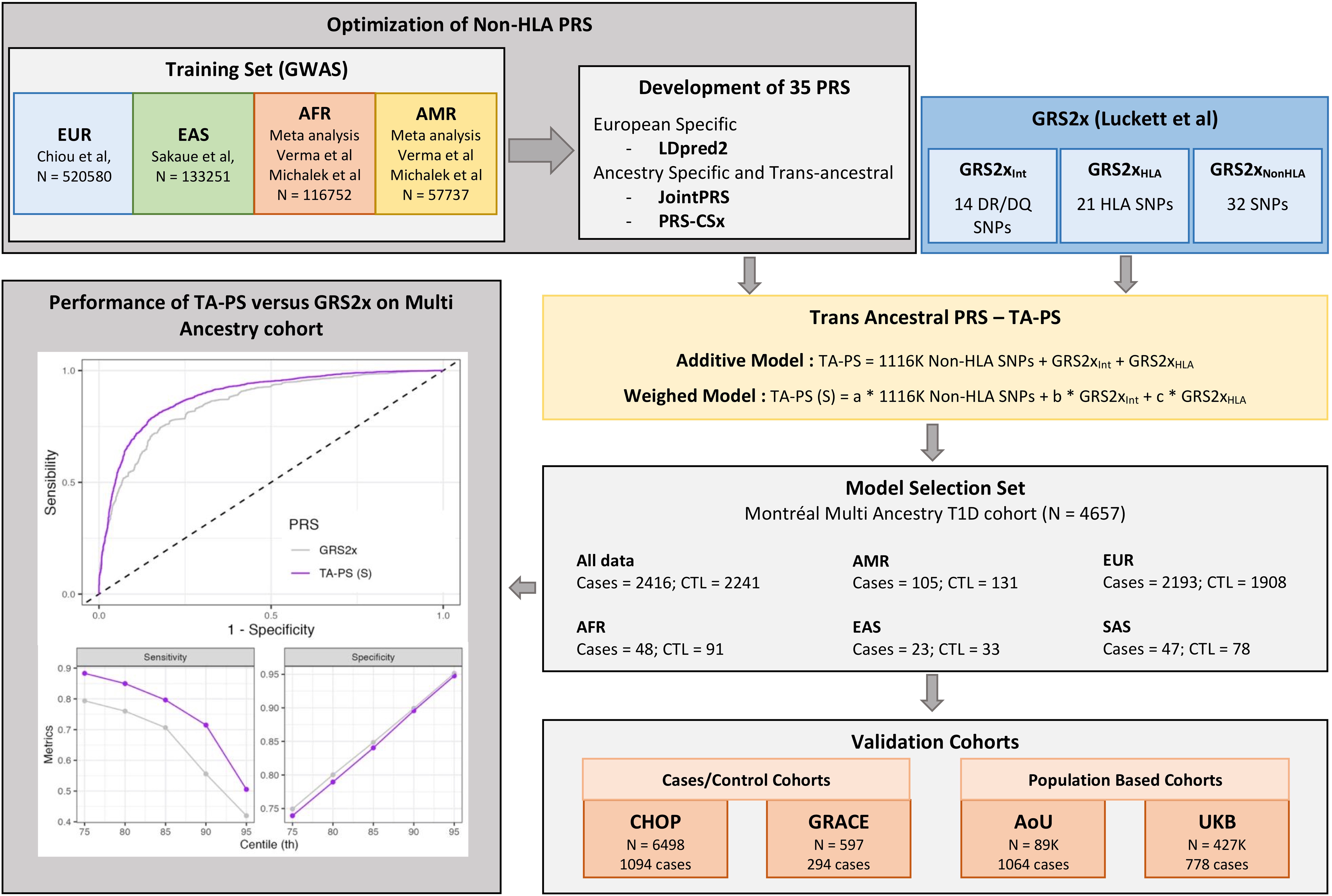
Flowchart with the design of our study.

## Results

### Testing of the PRS

First, we tested the performance of the 35 generated TA PRS (combined with the *HLA* part of GRS2x) by ancestry (**Figure 2, Supplementary Table 5**). For PRS-CSx, selection of phi values is presented in **Supplementary Note 3** (**Supplementary Figure 1, Supplementary Table 6**). As expected, both GRS2x and the newly built European ancestry-specific PRS using LDPred2 achieved higher AUROC in the European population compared to non-European populations. With multi-ancestral data incorporated, the TA scores consistently outperformed these two European-based PRS in the European (AUROC 0.88 for TA PRS vs 0.86 for GRS2x), African (AUROC 0.86 for TA PRS vs 0.83 for GRS2x), Hispanic (AUROC 0.86 for TA PRS vs 0.81 for GRS2x), East Asian (AUROC 0.82 for TA PRS vs 0.81 for GRS2x), and South Asian (AUROC 0.9 for TA PRS vs 0.87 for GRS2x) populations. Furthermore, the TA PRS outperformed or were comparable to the best-performing ancestry-specific PRS built using PRS-CSx or JointPRS. The results of the Delong test for comparison of the AUROC are shown in **Supplementary Table 7.**

**Figure 2.**
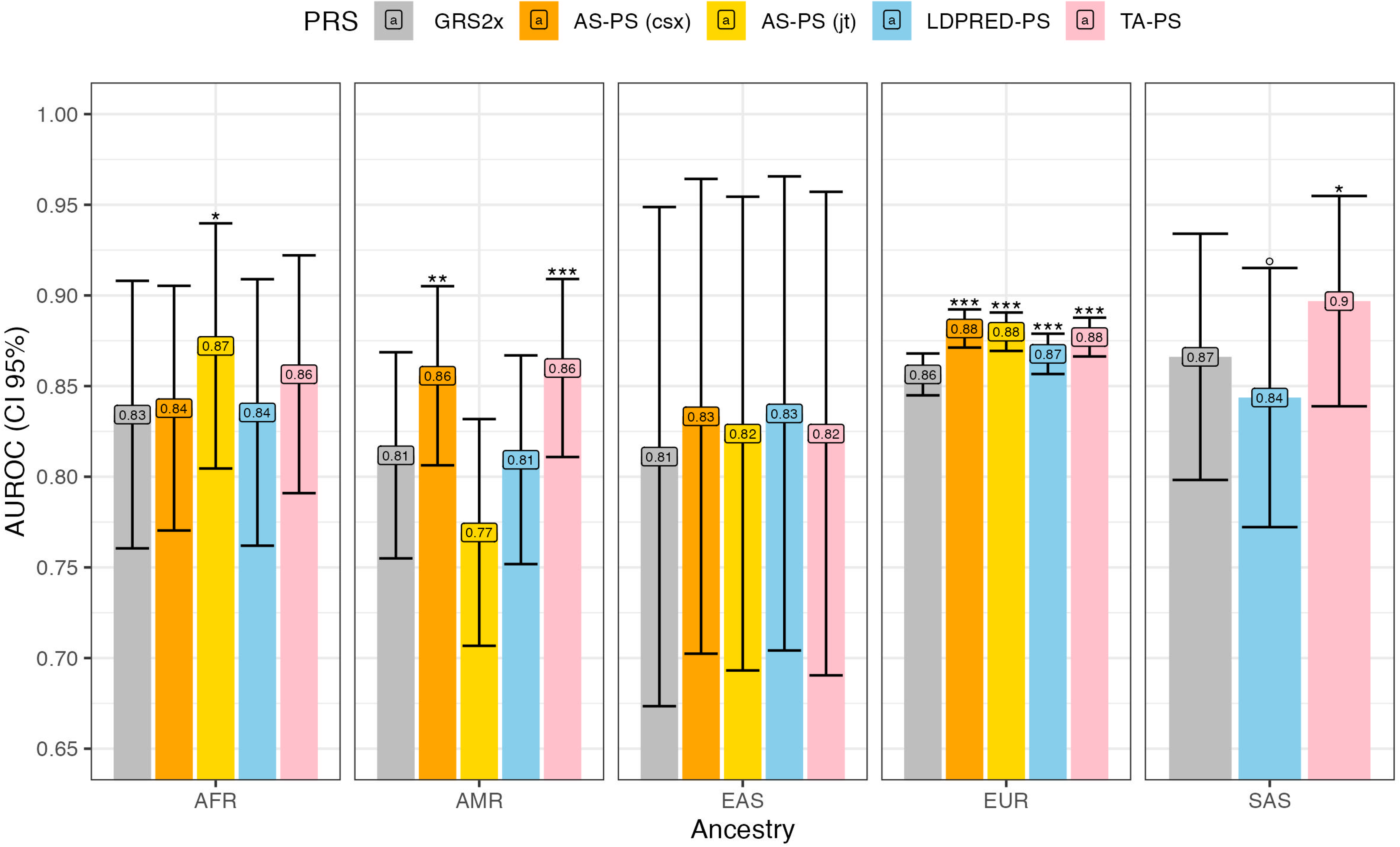
AUROC of the various PRS by ancestry and their 95% confidence intervals. AS-PS (csx) corresponds to the ancestry-specific PRS (one per ancestry). AS-PS (jt) JT corresponds to the ancestry-specific PRS (one per ancestry) developed with JointPRS. TA corresponds to the trans-ancestral PRS developed with PRS-CSx (phi = 1 x 10^-6^). All PRS are combine with the *HLA* part of GRS2x (GRS2x_Int_ + GRS2x_HLA_). Asterisks (*) represent the P-value of the Delong test between GRS2x and the different PRSs. Significance threshold: *P* < 0.001: ***; *P* < 0.01: **; *P* < 0.05: *; *P* < 0.10: °. MA: Multi-Ancestry.

### Selection of the best weighting approach for the PRS

We then focused on optimizing the combination of the TA scores with the *HLA* part of the GRS2x (GRS2x_Int_ and GRS2x_HLA_). **Figure 3** and **Supplementary Table 8** show the performance of the combined PRS using different weighting approaches and scaling methods (scale and MinMax), named TA-PS for additive model, TA-PS (S) for weighted model with scale transformation and TA-PS (M) for weighted model with MinMax transformation. The performance of the two versions of the weighted TA-PS in the European dataset (AUROC = 0.90) and of the TA-PS (S) in the Hispanic dataset (AUROC =0.86) was better than that of the unweighted version of TA-PS and of GRS2x. In the South Asian population, we found that only the crude TA-PS (AUROC= 0.90) performed better than GRS2x.

**Figure 3.**
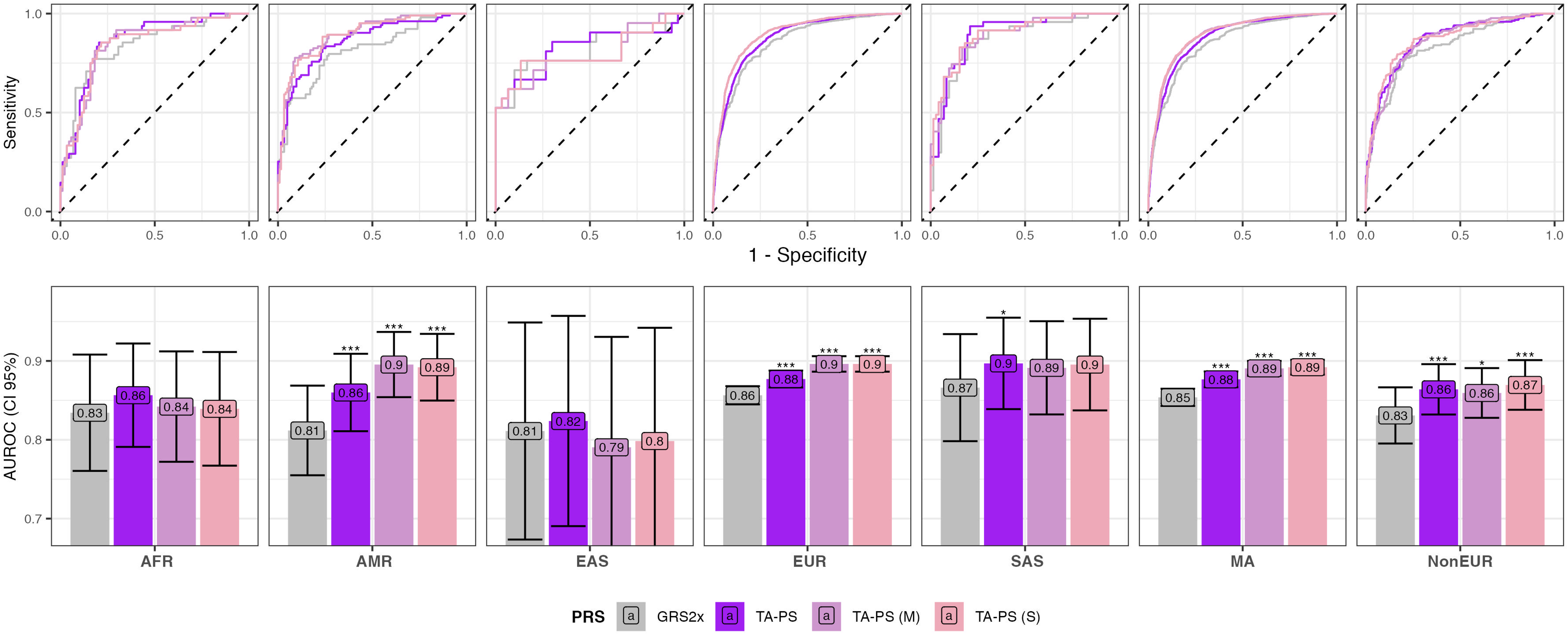
AUROC by ancestry of additive models versus weighted models. A) AUROC curve. B) AUROC and confidence interval (95%) by PRS and ancestry. TA-PS corresponds to the Trans-Ancestral score with phi = 10^-6^ with the full *HLA* part of the GRS2x (GRS2x_Int_ + GRS2x_HLA_). TA-PS (S) corresponds to the scaled weighted version while TA-PS (M) corresponds to the MinMax weighted version of TA-PS. Asterisks (*) represent the P-value of the Delong test between GRS2x and the different PRSs. Significance threshold: *P* < 0.001: ***; *P* < 0.01: **; *P* < 0.05: *; *P* < 0.10: °. MA: Multi-Ancestry.

For other ancestries, the small number of cases did not allow us to observe significant differences between the TA-PS in its weighted or crude versions and GRS2x. To increase the statistical power of our analysis, we conducted analyses on the entire multi-ancestry dataset (MA, N = 4657) and on the non-European dataset (NonEUR, N = 556) (**Supplementary Table 8**). In the multi-ancestry dataset (MA), GRS2x showed an AUROC of 0.85 while the TA-PS (S) obtained an AUROC of 0.89 (*P*_Delong_ = 4.67 x 10^-31^, **Supplementary Table 9**). In the Non-European dataset, GRS2x showed an AUROC of 0.83 versus an AUROC of 0.87 of the TA-PS (S) (*P*_Delong_ = 0.0007, **Supplementary Table 9**). Overall, these results suggest that weighting of the TA-PS can further improve type 1 diabetes prediction across ancestries on top of the crude TA-PS.

### Choice of the optimal threshold

We then compared the distribution of the TA-PS (in its crude or weighted versions) and GRS2x by ancestry. Specifically, we set a threshold at the 90^th^ centile in European controls for both scores and looked at their distribution in other ancestries (**Figure 4A**). For GRS2x, the 90^th^ threshold for the European dataset corresponds to the 86^th^ for the Hispanic dataset, to the 90^th^ for the East Asian and South Asian datasets, and to the 93^th^ for the African dataset. While for TA-PS (S), the 90^th^ threshold in the European population corresponds to the 83^th^ for the African, to the 85^th^ for the South Asian, to the 87^th^ for the East Asian and to the 91^th^ for the Hispanic populations.

**Figure 4.**
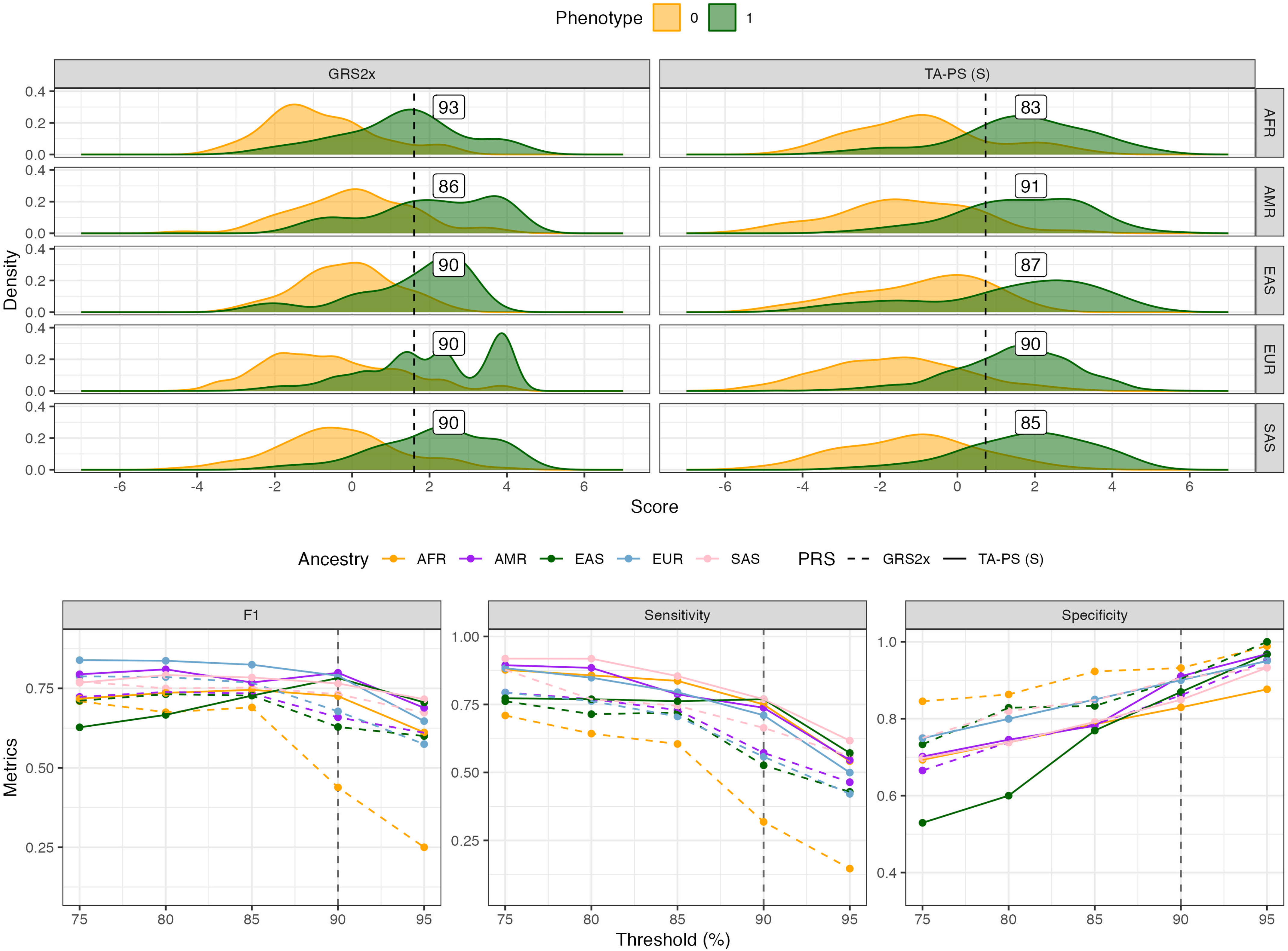
Distribution and performance of PRS by ancestry. A) Distribution of PRS by ancestry. The dashed line corresponds to the top 10% scores in controls of European ancestry (1.60 for GRS2x and 0.75 for TA-PS (S)). B) Performance metrics based on different thresholds determined in controls of EUR ancestry. MA: Multi-Ancestry.

We calculated, for each threshold determined in controls of European ancestry (95^th^, 90^th^,85^th^, 80^th^ and 75^th^ centile), different metrics (sensitivity (SEN), specificity (SPE) and F1) (**Figure 4B, Supplementary Table 10**). At the 90^th^ centile threshold, the average sensitivity per ancestry of the GRS2x was 0.53 (SEN_min_AFR_ = 0.32 and SEN_max_SAS_ = 0.66) while for TA-PS (S) it was 0.74 (SEN_min_EUR_ = 0.71 and SEN_max_SAS_ = 0.77). Regarding the specificity, it was higher on average for GRS2x (SPE_mean_AMR_ = 0.90, SPE_min_AMR_ = 0.86 and SPE_max_AMR_ = 0.93) than for TA-PS (S) (SPE_mean_AFR_ = 0.88, SPE_min_AFR_ = 0.83 and SPE_max_EAS_ = 0.91). As for the F1, it was on average higher for TA-PS (S) (F1_mean_AFR_ = 0.78, F1_min_AFR_ = 0.73 and F1_max_AMR_ = 0.80) than for GRS2x (F1_mean_AFR_ = 0.63, F1_min_AFR_ = 0.44 (AFR) and F1_max_SAS_ = 0.73).

At the 80^th^ percentile threshold, we found similar trends between GRS2x and TA-PS (S), notably that the average sensitivity was higher for TA-PS (S) (SEN_mean_ = 0.86) than for GRS2x (SEN_mean_ = 0.74). Similarly, the average F1 was 0.78 for TA-PS (S) and 0.74 for GRS2x.

In the entire multi-ancestry dataset (MA), at the 90^th^ threshold, the sensitivity of the TA-PS (S) was higher than that of GRS2x (SEN = 0.71 vs 0.56, *P*_z-test_ = 9.64 x 10^-51^, **Supplementary Table 11**) while the specificity was similar (SPE = 0.90, *P*_z-test_ = 0.57). In the Non-European subset, the sensitivity of the TA-PS (S) was higher than that of the GRS2x (SEN = 0.75 vs 0.53 p_z-test_ = 2.07 x 10^-10^), with comparable (*P*_z-test_ = 0.14) specificities (0.87 for TA-PS (S) vs 0.90 for GRS2x). For crude TA-PS, the metrics were overall better than those of GRS2x but slightly worse than those of its weighted version.

### Validation in independent cohorts

To validate the results obtained in our testing cohort, we applied GRS2x and TA-PS (S) in four independent cohorts, including two case/control studies (CHOP-CAG and GRACE) and two population-based studies (AoU and UKB) (**Figure 5, Supplementary Tables 12, 13, 14, Supplementary Figure 2**). We also included the results on the Montreal cohort in the figure for comparison.

**Figure 5.**
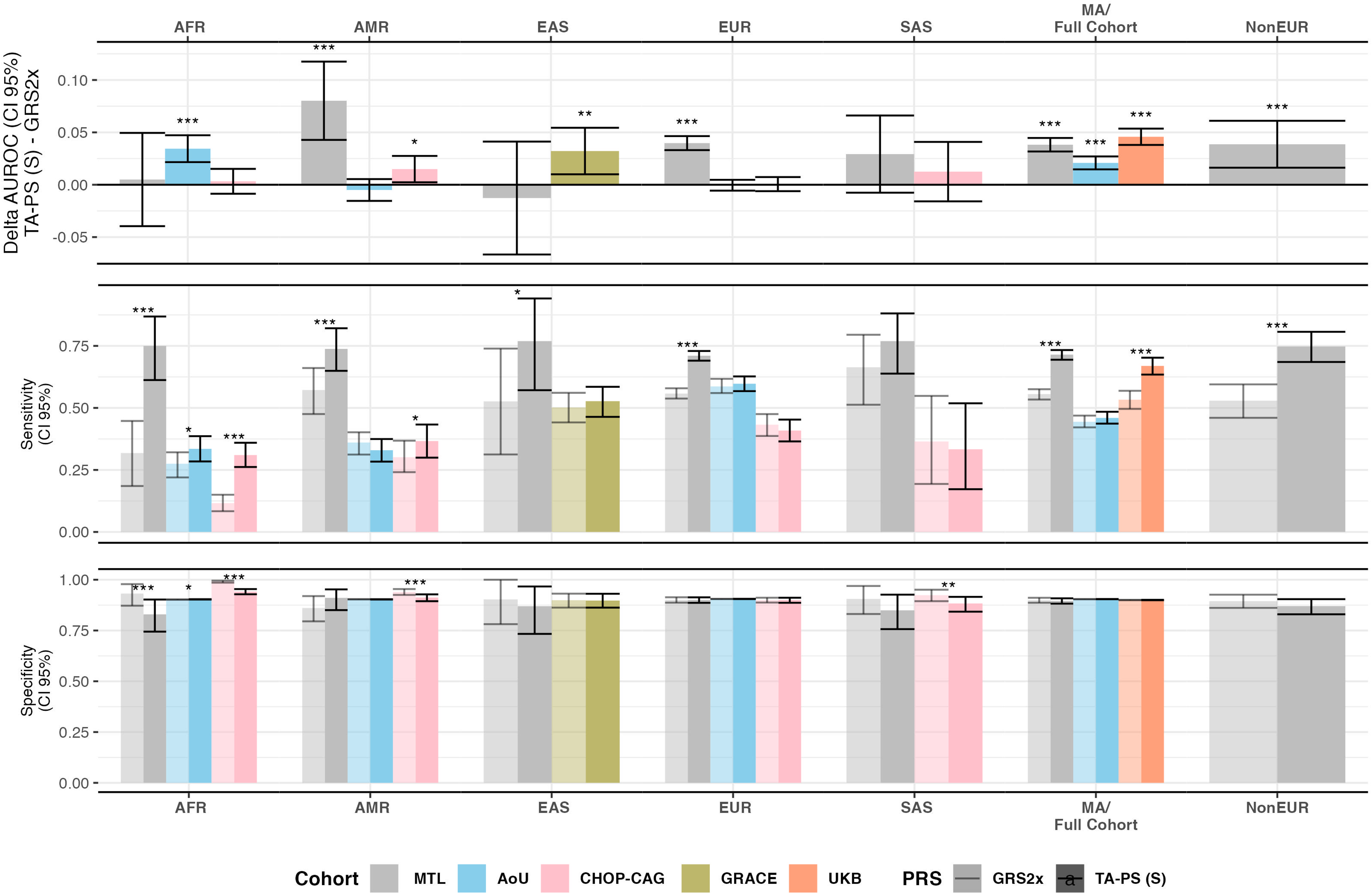
Performance of TA-PS (S) versus GRS2x by cohorts and ancestry. A) AUROC difference between GRS2x and TA-PS (S) and Delongs test (95% CI). Comparison between GRS2x and TA-PS (S) of sensitivity (B) and specificity (C) at the 10% threshold (90th percentile). Thresholds were estimated on controls for case/control cohorts and in the entire population for population-based cohorts. Confidence intervals (95% CI) were estimated by bootstrap and comparison was made with Z-test. Significance threshold: *P* < 0.001: ***; *P* < 0.01: **; *P* < 0.05: *; *P* < 0.10: °. MA: Multi-Ancestry.

In the East-Asian GRACE cohort, TA-PS (S) showed an improved AUROC compared to GRS2x (0.81 vs 0.78, *P*_Delong_ = 0.005). In terms of sensitivity, specificity and F1, there were no differences between the two PRS at the 90^th^ percentile threshold.

In the CHOP-CAG cohort, AUROC were comparable between GRS2x and TA-PS (S), except in admixed individuals where we found a significant difference (*P*_Delong_ = 0.02) in favour of the TA-PS (S). At the 90th percentile, for African subset, the sensitivity of TA-PS (S) was significantly higher than that of GRS2x (0.31 vs 0.12, *P*_z-test_ = 2.27 x 10^-30^) and conversely the specificity (0.99 vs 0.94) was higher for GRS2x compared to TA-PS (S) (*P*_z-test_ = 7.14 x 10^-84^), while remaining higher than 0.94 for both. Overall in African populations, the F1 score was higher with TA-PS (S) than with GRS2x (0.41 vs 0.20, *P*_z-test_ = 1.03 x 10^-14^). For the other ancestries we obtained comparable results between the two PRS.

Within AoU, the AUROC was higher for TA-PS (S) compared to GRS2x (0.75 vs 0.73, *P*_Delong_ = 3.65 x 10^-11^) with comparable sensitivity and specificity at the 90^th^ percentile. In the African subset, TA-PS (S) showed a better AUROC than GRS2x (0.65 vs 0.62 *P*_Delong_ = 1.46 x 10^-7^), and a better sensitivity (0.34 vs 0.28, *P*_z-test_ = 0.02) while maintaining an acceptable specificity (> 0.90).

In UKB (778 cases including 45 non-European), GRS2x obtained an AUROC of 0.81 which was lower than the AUROC of TA-PS (S) (AUC = 0.86, *P*_Delong_ = 8.72 x 10^-31^). At the 90^th^ percentile, the sensitivity of TA-PS (S) was better than that of GRS2x (0.67 vs 0.53, *P*_z-test_ = 2.56 x 10^-13^), with no significant differences in the specificity (SPE = 0.95, *P* > 0.90).

## Discussion

The objective of this study was to create an equitable genetic prediction tool for screening for type 1 diabetes in populations of diverse ancestral backgrounds. Building on previous developments in the field of type 1 diabetes genetic prediction in European populations (GRS2 and its updated version, GRS2x) [8,19], and recently available ancestry-specific GWAS datasets, totalling almost 30 thousand cases from various ancestries, we constructed our trans-ancestry PRS, named TA-PS. While this score did not focus on the optimization of the *HLA* component of the European scores, the non-*HLA* component was enriched with over a million SNPs, present in GWAS of four ancestries. The overall performance of TA-PS was better than that of GRS2x in our multi-ancestry testing cohort (N = 4657) based on metrics such as AUROC, sensitivity, specificity. Based on the AUROC, we observed a gain of 0.035 with the weighted and scaled TA-PS, compared to the GRS2x, in our entire multi-ancestry cohort. The 90^th^ centile of the distribution of this score showed, as a threshold to define high risk, an optimized sensitivity across all ancestries compared to the GRS2x, and a comparable specificity. Similar results were found in the European and Hispanic populations.

Our work enhances the use of type 1 diabetes PRS in population-wide screening programs. The importance to establish a common threshold using the same PRS, independently from the ancestral background, comes from the fact that a significant portion of the populations in many countries is admixed [1,11]. Adjustment for ancestry can be challenging, since reported ancestry can be inaccurate and participants in screening programs may refuse to disclose it [25]. Ancestry determined based on genetic data could be applied to select an ancestry-specific PRS, or an ancestry-specific threshold of a unique PRS, but clustering of admixed individuals within an ancestral group can still be challenging.

In our Montréal-based cohort, GRS2x did not achieve comparable sensitivity across ancestries at the 90^th^ percentile, particularly for samples of African ancestry, where the sensitivity was 0.31, while it was almost twice as high in participants of European ancestry (0.56). Contrarily, with TA-PS (S), we obtained higher sensitivities than GRS2x, but, importantly, this metric was more comparable among ancestries, ranging from 0.63 in East Asian to 0.79 in African populations, while maintaining specificities comparable to these of GRS2x, and greater than 0.84. When considering all non-European samples, at the 10% threshold, the sensitivity of TA-PS (S) was 0.72, which was significantly higher than that of GRS2x (0.53) while maintaining a specificity of 0.88, largely acceptable for a screening tool.

The validation in independent datasets confirmed the improved performance of the TA-PS over GRS2x: the multi-ancestry CHOP-CAG cohort and the East-Asian GRACE case/control studies demonstrated a gain in sensitivity of TA-PS (S) compared to GRS2x using as threshold the top 10% of the distribution. The AoU population cohort also demonstrated a significant gain in terms of AUROC using the TA-PS (S) across its entire multi-ancestry dataset but also specifically for African Americans. Furthermore, within this African subset, a gain in terms of sensitivity at the 90^th^ percentile threshold was observed. Finally, in UKB, we confirmed the TA-PS (S) outperformed GRS2x both in terms of AUROC and sensitivity (at the 90th percentile) while maintaining a specificity equal to 0.95. All these results support that TA-PS (S) allows to increase the prediction performance compared to GRS2x but also allows to increase the sensitivity while maintaining an acceptable specificity, at the top 10% of the distribution threshold used in previous studies.

Regarding the limitations of our study, the most important is absence of specific GWAS data for certain ancestries, notably South Asians or Indigenous peoples, limiting optimisation of the genetic predictors in these ancestries. The fact that we only optimized the non-*HLA* part of the score is another limitation, which should be addressed in future studies, as the *HLA* loci can explain at least 50% of the heritability of T1D. In order to optimize the *HLA* component using a similar approach as that of the GRS2 or GRS2x [8,19], access to large, ancestry-specific whole genome sequence data is necessary, which are currently lacking.

Also, it is notable that a previous study reported ancestry-specific PRS with higher performance metrics than TA-PS (S). Specifically, an African-specific PRS developed and tested in the CHOP-CAG cohort obtained an AUROC of 0.83 which is higher than that of our trans-ancestral score [10], but the testing cohort was not exactly the same as our CHOP-CAG validation dataset, and external validation was lacking. While the enhanced results obtained by TA-PS (S) in the European dataset of our testing cohort may reflect some degree of overfitting, given that the weights were estimated on the same samples, it nonetheless showed consistent performance in the independent, predominantly European UKB. The AUROC obtained with GRS2x and with TA-PS in the UKB and AoU were lower than those reported using the GRS2x or GRS2 in their respective publications [8,19]. This discrepancy can be explained by differences in the definitions of cases and controls in these biobanks. For instance, under previous definitions requiring diagnosis before a certain age threshold, the proportion of cases in UKB was 0.11% (408 cases and 365,622 controls) [19], while with our definition, the proportion was 0.18% (778 cases and 427,069 controls). In AoU, using previous definitions, the proportion of cases was 0.04% (147 cases and 317,817 controls), while we obtained a proportion of 1.12% (1,064 cases and 88,635 controls) following Szczerbinski et al, who applied stricter criteria for healthy controls and excluded individuals who did not clearly meet the case or control definitions [26]. Importantly, when evaluated on the same population of cases and controls, TA-PS outperformed or was at least comparable to GRS2x. Notably, our validation cohorts were either case/control, or population-based biobanks (AoU or UKB) requiring voluntary participation, which are not necessarily representative of the general population.

Data from prospective population-wide screening programs, such as the Canadian CanScreenT1D research consortium (https://canscreent1d.ca/), will be necessary to validate TA-PS in future studies.

To conclude, we constructed a trans-ancestry score for type 1 diabetes, named TA-PS, based on the largest and most recent GWAS datasets and employing the widely used PRS-CSx method for trans-ancestral PRS construction. TA-PS, in its scaled version, when applied in five independent case/control or population-based cohorts, yielded more comparable results among ancestries in terms of prediction accuracy and sensitivity than the European-based GRS2x. Future efforts, leveraging large-scale sequencing data from type 1 diabetes cases and controls from various ancestries, should focus on the optimization of the *HLA* part of this score.

## Data availability

The list of GWAS used and full summary statistics available are presented in the supplementary material (Supplementary Table 1).

## Code availability

R codes used to generate the PRS are available in Zenodo.

## Acknowledgements

We thank all participants in the GWAS consortia, data from which were used in this study. This work was funded by the Breakthrough T1D international (grant key: 2-SRA-2022-1262-S-B). D.M. is a Fonds de Recherche du Quebec-Santé (FRQS) Junior 1 Scholar and has received a Career Development Award from ENRICH (Empowering Next-Generation Researchers in Perinatal and Child Health). G.B.L is a Fonds de Recherche du Quebec-Santé (FRQS) Junior 1 Scholar. We gratefully acknowledge All of Us participants for their contributions, without whom this research would not have been possible. We also thank the National Institutes of Health’s All of Us Research Program for making available the participant data examined in this study. For UKB data were accessed under the project numbers 399545. Funding for the GRACE cohort: Key R&D Program Projects in Zhejiang Province (2024C03154) and with ethics approval No. is 2024-IRB-0270-P-0.

## Author contributions

D.M., C.G., C.P. conceived the study and supervised the analyses. B.J. drafted the manuscript, developed the PRS and performed the analyses on the Montreal based cohort. H.Q. performed the analysis on CHOP-CAG cohort. T.L. performed the analysis on All of Us biobank. W.B. performed the analysis on UK Biobank. K.L. and Y.L. performed the analysis on GRACE cohort. All authors contributed in study design, reviewing and writing the manuscript. All authors critically reviewed and approved the final version of the manuscript.

