## Supplemental Figure 1 for "Development and validation of a Trans-Ancestry polygenic risk score for Type 1 Diabetes"

**Supplementary Note 1**

To increase statistical power and expand the ancestral diversity of our study, we incorporated recently published type 1 diabetes GWAS datasets from individuals of African ancestry (AFR) and the Hispanic (AMR) population. We conducted separate meta-analyses of GWAS summary statistics using the sample-size weighted fixed-effects model implemented in Metal, which allows for the combination of studies with different measurement scales. The first meta-analysis included summary statistics from individuals of African ancestry (($N_{AFR\_Michalek}$ = 891, $N_{AFR\_Verma}$ = 115861), while the second focused on those from the Hispanic population ($N_{AMR\_Michalek}$ = 308, $N_{AMR\_Verma}$ = 57429) [1,2].

The PRS-CSx method was applied using the python package available on Github (getian107/PRS-CSx) which allows to estimate posterior SNP effect sizes inferred according to continuous coupled shrinkage (CS) priors between populations. PRS-CSx offers LD reference panels from UKB or 1K Genomes Project, but to avoid overfitting with our controls, we chose to use the UKB reference panel. PRS-CSx requires a shrinkage parameter. We tried a set of different shrinkage parameters (phi = $10^{-2}$; $10^{-4}$; $10^{-6}$; $10^{-8}$; $10^{-10}$; $10^{-12}$) and selected the one yielding the scores with the best prediction performance. PRS-CSx generated a set of ancestry specific PRS (AS, at a range of phi = $10^{-2}$; $10^{-4}$; $10^{-6}$; $10^{-8}$; $10^{-10}$; $10^{-12}$) for AFR, AMR, EAS and EUR, as well as a set of Trans Ancestry PRS (TA, at a range of phi = $10^{-2}$; $10^{-4}$; $10^{-6}$; $10^{-8}$; $10^{-10}$; $10^{-12}$).

The JointPRS method is similar to PRS-CSx but uses a slightly different algorithm which also allows the estimation of posterior effect sizes of SNPs from multi-ancestry GWAS and LD reference panels. The python package is available on Github (LeqiXu/JointPRS). In the absence of tuning data, we ran JointPRS in its "JointPRS-auto" version. Similarly, to PRS-CSx, we used the LD reference panels from UKB. JointPRS generated only ancestry- specific PRS for AFR, AMR, EAS and EUR populations.

Finally, we constructed an enhanced European PRS using LDpred2 to optimize the non-HLA part of the GRS2x. To do this, we used as reference the GWAS by Chiou et al, which is the largest European GWAS to date. We conducted LDpred2 using SNPs present in HapMap3+ and in total, 1,422,515 SNPs matched between the reference GWAS and HapMap3+.

**Supplementary Note 2**

We then tested the performance of the various PRS in independent case/control or population-based cohorts of various ancestries (**Supplementary Table 4**). Specifically, in the multi-ancestry All of Us Biobank (AoU, N = 89,699), in a case-control multi-ancestry cohort from the Children's Hospital of Philadelphia (CHOP-CAG, N = 6498), in the UK Biobank (UKB, N = 427,847) and in a Chinese case/control cohort set up in pediatric centers named Genetic Risk Assessment for Chinese Eaglet-T1D (GRACE, N = 597).

Within AoU ($N_{cases}$ = 1084; $N_{CTL}$ = 88,635), type 1 diabetes diagnosis (T1D-EHR) was defined according to previously published criteria [3] and cases of undefined diabetes were excluded from the controls. More precisely, the cases were defined as follows: At least 1 ICD-9, ICD-10, or SNOMED code of type 1 diabetes in EHR data, must have been prescribed insulin, must not have been prescribed any other diabetes medication besides insulin and no diagnosis of cancer, cystic fibrosis, drug-induced diabetes, maturity-onset diabetes of the young (MODY), gestational diabetes mellitus (GDM), or post-transplant diabetes. While the controls were defined as follows: must have EHR data, no EHR record of any type of diabetes, must have at least 1 measurement of fasting glucose, random glucose, or HbA1c, no record of fasting glucose ≥100mg/dl, random glucose ≥200mg/dl, or HbA1c ≥5.7%, no self-reported medical history of any type of diabetes, must not have been prescribed any other diabetes medication, no diagnosis of cancer or cystic fibrosis.

In UKB ($N_{cases}$ = 778; $N_{CTL}$ = 427,069), the type 1 diabetes definition was made according to following criteria: insulin-dependent diabetes (ICD-10), and reported age <25 years at diagnosis. For controls we excluded type 2 diabetes and undefined diabetes.

In the CHOP-CAG ($N_{cases}$ = 1094; $N_{CTL}$ = 5404) and GRACE cohorts ($N_{cases}$ = 294; $N_{CTL}$ = 303), the diagnosis of type 1 diabetes was made by clinicians and retrieved from patient hospital records.

**Supplementary Note 3**

Regarding PRS-CSx we tried a set of shrinkage parameters (phi), the results obtained are presented in **Supplementary Table 6** and Supplementary **Figure 1**. In the main manuscript, TA-PS refers to the trans-ancestry score developed with PRS-CSx with phi = 10-6. This phi value, among all generated TA scores, is the one that performs the best across all ancestries ($AUROC_{mean}$ = 0.86, $AUC_{sd}$ = 0.03, $AUROC_{min}$ = 0.82 (EAS) and $AUROC_{max}$ = 0.90 (SAS).

**Supplementary Figures**

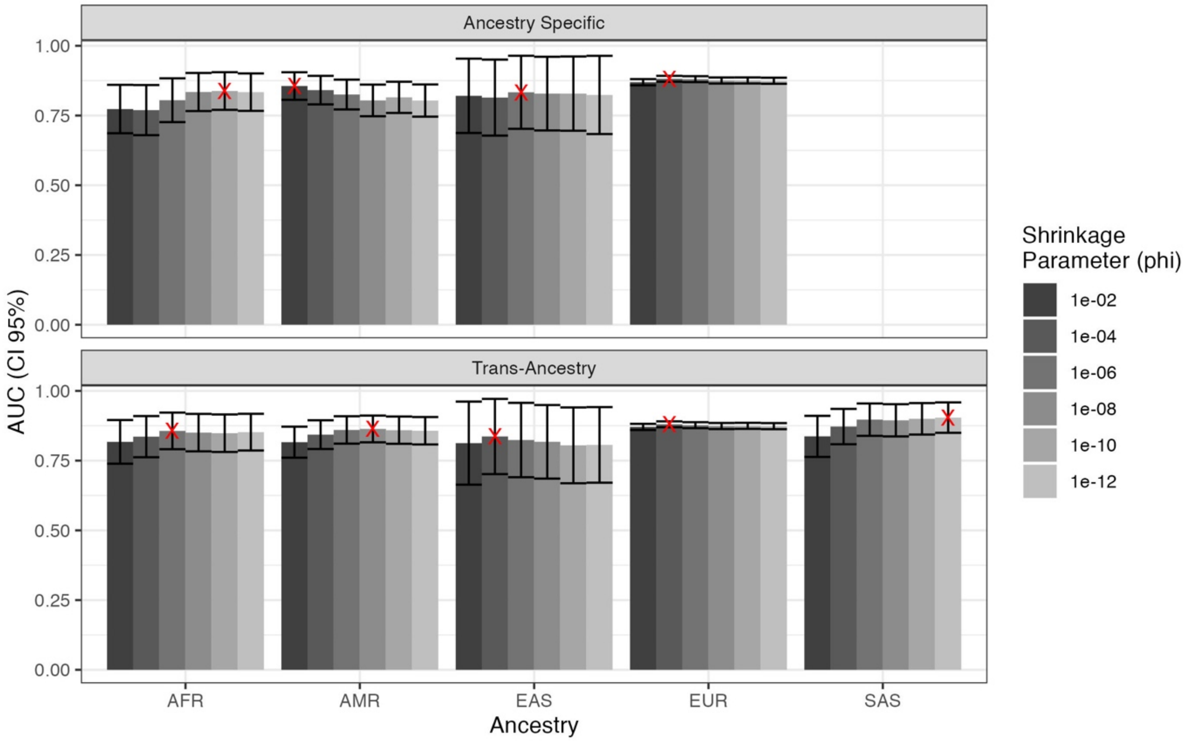


**Supplementary Figure 1.** AUROC of the various PRS by ancestry and their 95% confidence intervals. Set of PRSs developed with PRS-CSx combined with the HLA portion of GRS2x. Six shrinkage parameters (phi) tested for each ancestry-specific PRS or for the trans-ancestry PRS. The red cross marks the best PRS.

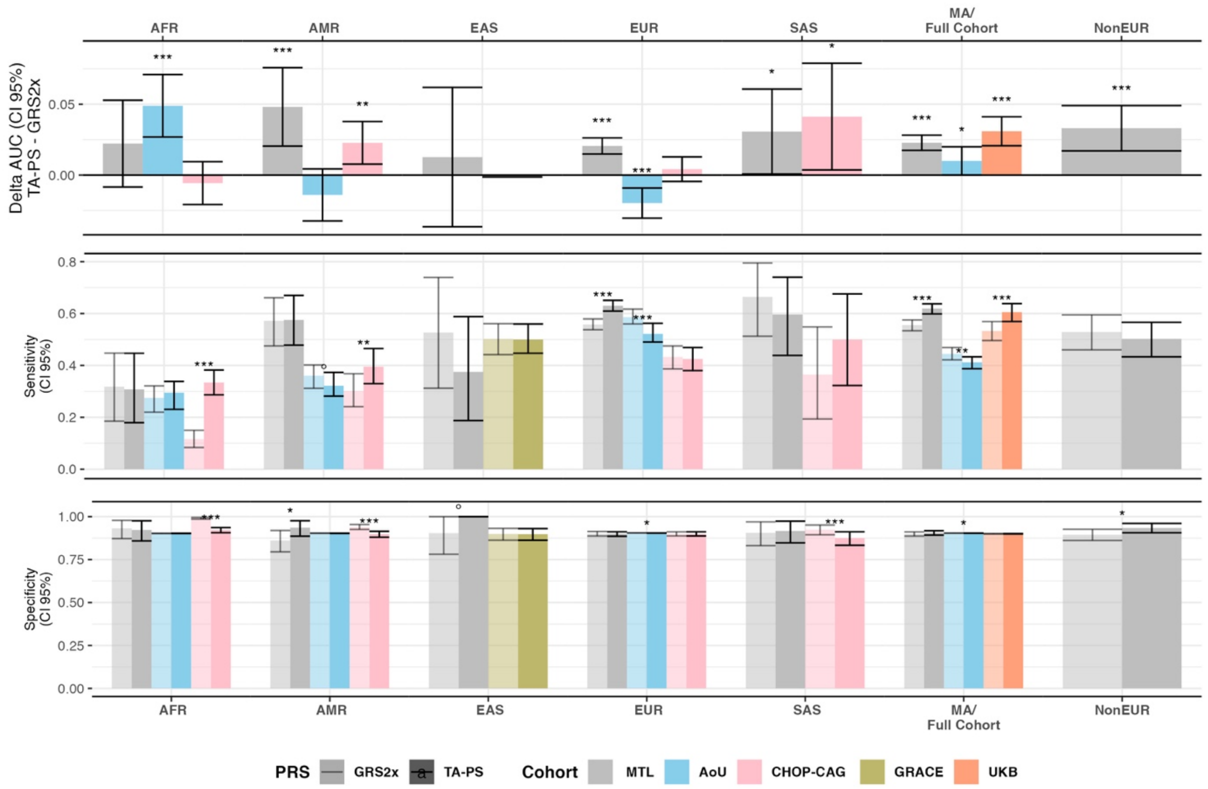


**Supplementary Figure 2.** Performance of TA-PS versus GRS2x by cohorts and ancestry. Upper panel: AUC difference between GRS2x and TA-PS and Delongs test (95% CI). Comparison between GRS2x and TA-PS of sensitivity (center panel) and specificity (bottom panel) at the 10% threshold (90th percentile). Threshold estimated on controls for case/control cohorts and for population cohorts the threshold was estimated on the whole population. Confidence intervals (95% CI) estimated by bootstrap and comparison made with Z-test. Significance threshold: p < 0.001: ***; p < 0.01: **; p < 0.05: *; p < 0.10: °. MA for Multi-Ancestry.

<u>References</u>

1. Michalek, Dominika A, Courtney Tern, Wei Zhou, Catherine C Robertson, Emily Farber, Paul Campolieto, Wei-Min Chen, Suna Onengut-Gumuscu, et Stephen S Rich. 2024. «

A multi-ancestry genome-wide association study in type 1 diabetes ». Human Molecular Genetics 33 (11): 958-68. https://doi.org/10.1093/hmg/ddae024.

2. Szczerbinski, Lukasz, Ravi Mandla, Philip Schroeder, Bianca C. Porneala, Josephine H. Li, Jose C. Florez, Josep M. Mercader, Miriam S. Udler, et Alisa K. Manning. 2024. « Algorithms for the Identification of Prevalent Diabetes in the All of Us Research Program Validated Using Polygenic Scores ». Scientific Reports 14 (1): 26895.
3. Verma, Anurag, Jennifer E. Huffman, Alex Rodriguez, Mitchell Conery, Molei Liu, Yuk-Lam Ho, Youngdae Kim, et al. 2024. « Diversity and Scale: Genetic Architecture of 2068 Traits in the VA Million Veteran Program ». Science 385 (6706): eadj1182.
